# Forecast of the covid19 epidemic in France

**DOI:** 10.1101/2021.04.13.21255418

**Authors:** Loïc Pottier

**Affiliations:** Education nationale

## Abstract

With a mathematical method based on linear algebra, from open access data (data.gouv.fr, google, apple) we produce forecasts for the number of patients in intensive care in France with an average error of 4% at 7 days, 7% at 14 days, 8% at 21 days, 10% at one month, 17% at 2 months, and 31% at 3 months. For the other epidemic indicators, the error is on average 6% at 7 days and 25% at 2 months.

## 1 Introduction

The method we use begins with the computation of maximum correlations between the data of the daily indicators of the epidemic (hospitalizations, intensive care, number of cases, deaths, etc.) and those, arbitrarily offset in time, of the data of its context (holidays, weather, mobility of people, curfew, etc.), for each day and each French department.

We deduce from this the time offsets between contexts and indicators, for example 19 days between attendance at workplaces (context) and the reproduction rate ^1^ associated with the number of patients in intensive care (indicator).

Then, from these offsets, a linear approximation by quadratic optimization and forecast of the reproduction rates is computed. Approximations of indicators whose effective reproduction rate is approximated with an average error greater than 5% are rejected. For the others, we deduce the corresponding indicators from the approximations and forecasts.

This is done for each department (for departments where the full data of the day is present at the time of the calculation, i.e. generally around 88).

At the end of this article, we present the current forecasts for the number of patients in intensive care. Detailed and updated daily results can be found here: https://cp.lpmib.fr/medias/covid19/synthese.html.

The only assumptions which are made are that the data of the context keep in the future the values which they have at the present day, except for the weather forecast, where one takes the values of the past year at the same time, and for school holidays, which have been planned for a long time.

## 2 Correlations and offsets

The data relate to the epidemic indicators (emergencies, intensive care, deaths, positive tests, etc.) and the contexts (weather data: temperature, pressure, mobility data provided by google: frequentation of shops and places of leisure, workplaces, etc.). These are daily data (not cumulative). They appear, for each data *x* ∈ {1, …, *N*} (indicator or context), and each department *d* ∈ {1, …, *D*}, as a vector of values: 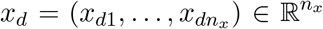, corresponding to an interval of *n*_*x*_ days [*j*_0_(*x*), *j*_0_(*x*) + *n*_*x*_[.

We use *N* = 48 data, which concerns *D* = 86 departments and more than 464 days.

We start by calculating the correlation coefficients between two data items *x* and *y*, for all the time offsets *t* of at most *t*_*max*_ = 40 days.

For each department *d*, we calculate the average value of *x*_*d*_

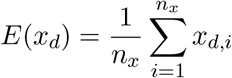

then we complete by the value 0 the vector *x*_*d*_ − *E*(*x*_*d*_) for the days when it is not defined in the interval

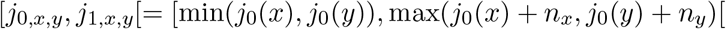

We then obtain the vector *x*^*′*^_*d*_. Likewise for *y*. We then define *x*_*f*_ by concatenation of the values for each department:

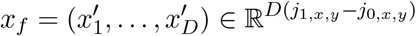

and *y*_*f*_ idem.

We define the offset of *t* of a vector *z* ∈ℝ^*n*^ by

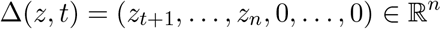

where we therefore complete by *t* zero values.

The correlation coefficient between *x* and *y* shifted by *t* is then defined by

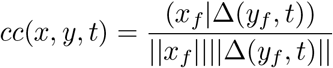

where (.|.) And ||.|| denote the euclidean product and norm.

The correlation coefficient from *x* to *y* is then

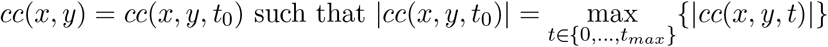

and we define the offset from *x* to *y* by

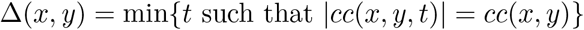

For example, if we note *work* the frequentation of workplaces (data from Google) and *Rintensivecare* the reproduction rate associated with the number of patients in intensive care (data from Santé Publique France), we obtain the following offsets in days:

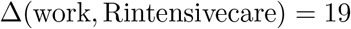

and

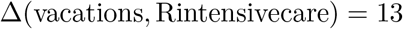

And we have the correlation coefficients

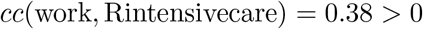

and

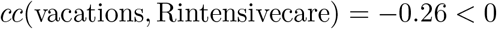

which suggests possible causalities: an increase in work place attendance seems to cause an acceleration of intensive care 19 days later, and a period of school vacations seems to cause a slowdown in intensive care 13 days later.

We then define the dependencies of a data *y* as

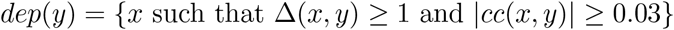

The value 0.03 is low and minimizes a posteriori the error of the forecasts, but has little influence on them.

These dependencies with their offsets will make it possible to predict an indicator of the epidemic, first by predicting its effective reproduction rate, then by deducing its values, as explained below.

## 3 Linear forecast coefficients

Now we have day offsets between some of the data. We will say that a data *y* depends on a data *x* if *x* ∈ *dep*(*y*), i.e. if we obtained, in the previous step, a offset Δ(*x, y*) *>* 0 from *x* to *y* and a sufficient *cc*(*x, y*) correlation. We then consider that a data *y* over a period of time [*j*_0_, *j*_1_] will depend on the values of the data *x*_*i*_ ∈ *dep*(*y*) over the periods [*j*_0_ − Δ(*x*_*i*_, *y*), *j*_1_ − Δ(*x*_*i*_, *y*)].

Let’s fix a department. Let us call *A* the matrix whose column *i* is formed by the values of *x*_*i*_ in this department over the period [*j*_0_ − Δ(*x*_*i*_, *y*), *j*_1_ − Δ(*x*_*i*_, *y*)], and *B* the column vector formed by the values of *y* in this department over the period [*j*_0_, *j*_1_].

We would like to find a family of coefficients *C* such that *AC* = *B*. But there is usually no solution for this equation, because *A* has more rows (days) than columns (the data *x*_*i*_ on which *y* depends). We then try to minimize

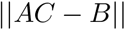

This is a convex quadratic problem, the solution of which is simply obtained with

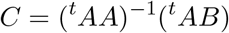

(if ^*t*^*AA* is invertible, which is the case in practice).

Indeed, if *f* (*X*) = ||*AX* − *B*||^2^, then

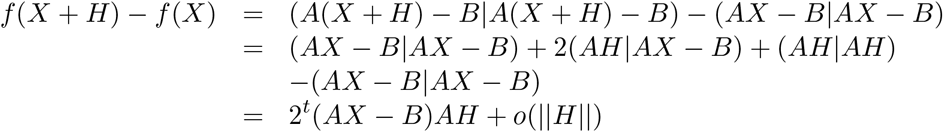

So the differential of *f* in *X* is *Df* (*X*) : *H 1*→ 2^*t*^(*AX* − *B*)*AH*. It is clear that *f* is convex, so it is minimal when its differential is zero, i.e. when ^*t*^(*AX* − *B*)*A* = 0, i.e. when ^*t*^*AAX* =^*t*^ *AB*, therefore, if ^*t*^*AA* is invertible, when *X* = (^*t*^*AA*)^−1^(^*t*^*AB*).

With *C* we can predict a value for the data *y* on the day *j*_1_ + 1, simply by computing *XC*, where *X* is the row vector of the values of *x*_*i*_ for the days *j*_1_ − Δ(*x*_*i*_, *y*) + 1:

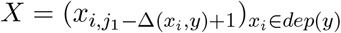

We then provide values for all data for one day in parallel, then the next day, etc. If a data is not predictable (because it has no dependency, or ^*t*^*AA* is not invertible), we keep its previous value. We do this for each department.

This makes it possible to predict values for data in the future, if we assume the data of the contexts to be constant from the present for the future, except for the meteorological data that we take from the past year to the same date, and vacation dates, which are known for the future.

## 4 Effective reproduction rate

The visible effects of the epidemic are measured with the indicators of the health system, and are essentially the result of contamination between a sick person and a healthy person. The *R* reproduction rate is difficult to determine because it changes every day, and not all who are sick and who infects them are known.

To approximate it, we will determine for each indicator and each day of the epidemic an effective reproduction rate that we will note again *R*, using an estimated serial interval *s* = 4.11 (this is the average number of days between two successive contaminations in a contamination chain). If the chosen epidemic indicator is given each day by a function *f* (for example the daily number of new hospitalizations), then its *R* checks

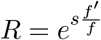

so that *f* (*x* + *s*) = *R*(*x*)*f* (*x*).

We obtain this expression simply by considering *f* as locally exponential, which is the case during an epidemic: assuming that a patient infects *α* people on average in one day, we have, for any instant, *x*,

*f* (*x* + 1) = *αf* (*x*), so *f* (*x* + 1) − *f* (*x*) = (*α* − 1)*f* (*x*),

therefore, by identifying the derivative and the discrete derivative, *f′* = (*α* − 1)*f*, therefore 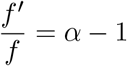,

then by integration we obtain ln *f* (*x*) = (*α* − 1)*x* + *c*, so *f* (*x*) = *e*^(*α*−1)*x*^*f* (0), so *f* (*x* + *s*) = *e*^(*α*−1)*s*^ *f* (*x*) and finally 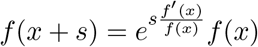,

hence 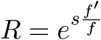.

The value *R*(*x*) of the reproduction rate in fact varies every day *x*: it is maximum at the start of the epidemic, and then decreases to 0, when the population is immune and the epidemic subsides.

Note that the discrete derivative

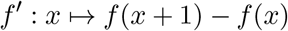

is in practice computed on the smoothed values 2 times over 7 days.

In the continuous case, we can find the value of the indicator at day *j* from the function *R* as follows:

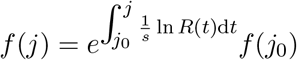

We adapt this formula to the discrete case by discrete integration and then global correction for the part corresponding to the real data (essentially we normalize to obtain the real integral of *f* on the past and its correct value in the present).

## 5 Summary of the method

To summarize, the indicator prediction process is therefore as follows:

1. computation of the effective daily *R* reproduction rates of the indicators.
2. computation of correlations and offsets between contexts and effective reproduction rates of indicators
3. we deduce the contexts on which these rates depend.
4. from these contexts, compute the linear forecast coefficients of the rates *R*.
5. with the linear forecast coefficients, computing forecasts of *R* rates in the future.
6. by discrete integration and normalization on the past, computation of forecast indicators in the future.

## 6 Results and evaluation

To evaluate the method, we apply it in the past. We use the linear forecast coefficients computed over the period of the epidemic (from April 2020 to April 2021) to forecast the values of the indicators 7 days after, for example, on December 1, 2020, then 14 days after, etc., until 3 months later (March 1). Then the relative errors between the predicted values and the actual values are computed.

On April 12, 2021, the average relative error between the actual values and the values forecast for 4 months is less than

- at 7 days: 4 % for intensive care (6% for all indicators)
- 14 days: 6 % (8 %)
- 28 days: 10 % (11 %)
- 2 months: 16 % (32 %)
- 3 months: 22 % (46 %).

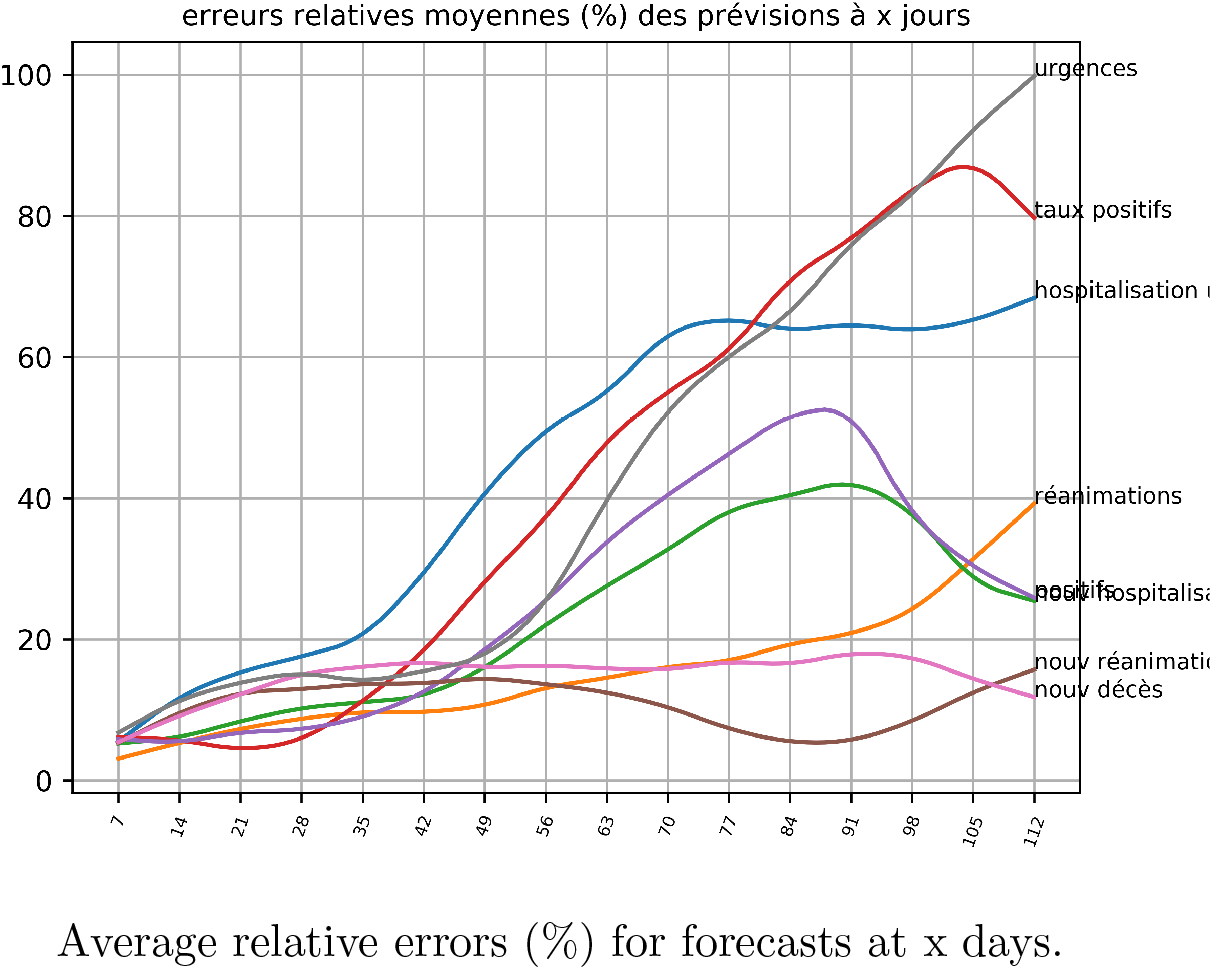

The peak of the number of intensive care of the current wave is thus foreseeable for the end of April, with a plateau, without knowing what will happen next:

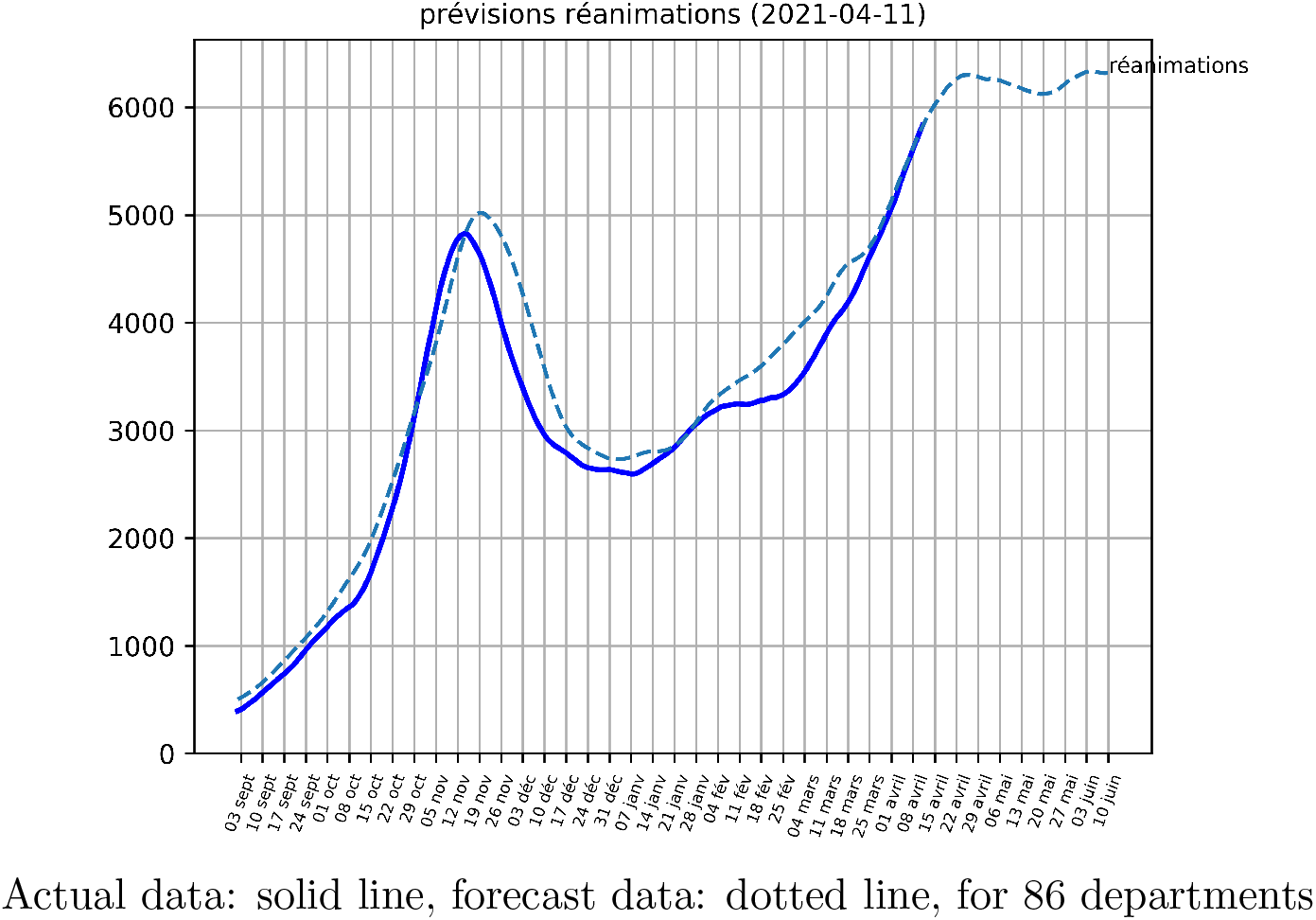

Using the forecast coefficients computed from the data before the start of the forecast, we obtain, for forecasts over 2 months for the last 42 days:

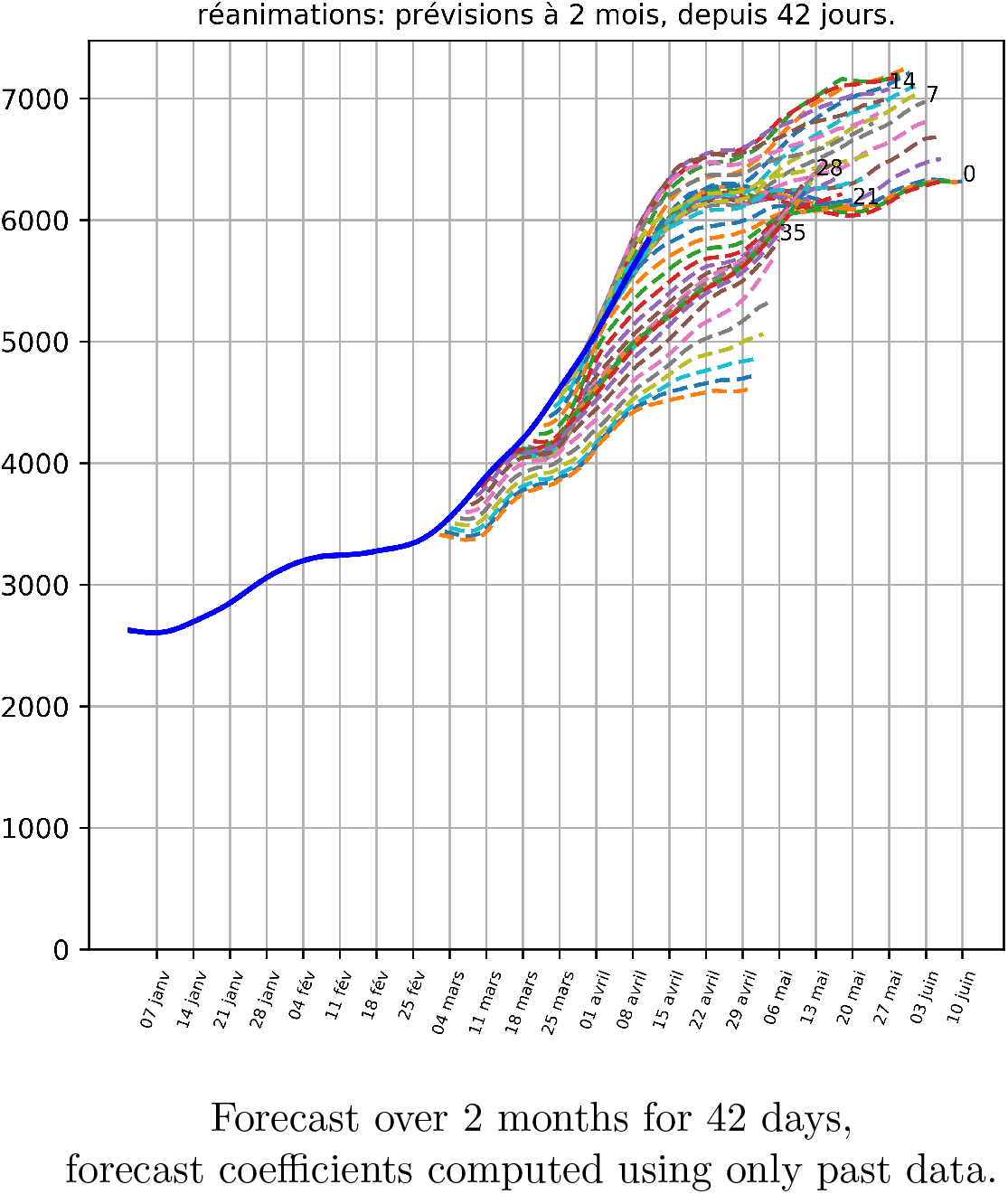

We can also evaluate the method by comparing it with a linear approximation (by tangent to the curve) or quadratic (use of the first and second derivatives in the present to approximate the curve by a polynomial d th degree 2). These approximations are always less good, with errors of 8 % at 7 days, 17 and 21 % at 14 days, and more than 50 % from 50 days.

We can also compare with the short-term forecasts of [1]: they give an error of 6 % to 7 days and 11 % to 14 days for critical care beds.

Full forecasts are updated daily on the web page https://cp.lpmib.fr/medias/covid19/ _synthese.html

The code can be found here: https://github.com/loicpottier/covid19

## Data Availability

Data are open access data from data.gouv.fr, google, apple.

https://www.data.gouv.fr/fr/

https://www.google.com/covid19/mobility/

https://covid19.apple.com/mobility

## Footnotes

1 the effective reproduction rate is the average number of people infected by a patient (if below 1, the epidemic decreases, if above it increases). It is noted effective R, Reff, R0 or R in the literature.

